# Association of Extent of Resection and Functional Outcomes in Diffuse Low-Grade Glioma: Systematic Review & Meta-Analysis

**DOI:** 10.1101/2022.09.25.22280331

**Authors:** Mustafa Elsheikh, Elsie Bridgman, Jose Pedro Lavrador, Simon Lammy, Michael Tin Chung Poon

## Abstract

**Background:** Surgical resection offers survival benefits in patients with diffuse low-grade glioma (DLGG) but its association with functional outcomes is uncertain. This systematic review assessed functional outcomes associated with extent of resection (EoR) in adults with DLGG.

**Methods:** We searched Medline, Embase and CENTRAL on the 19^th^ of February 2021 for observational studies reporting functional outcomes after surgical resection for patients aged ≥18 years with a new diagnosis of supratentorial DLGG according to any World Health Organization classification of primary brain tumors. The Newcastle-Ottawa Scale (NOS) informed our risk of bias assessments. The proportion of patients returning to work within 12 months entered a random-effects meta-analysis. PROSPERO registration number CRD42021238387.

**Results:** There were seven eligible moderate to high-quality (NOS >6) observational studies identified from 1,183 records involving 234 patients with DLGG. Functional outcomes reported included neurocognition (n=2 studies), performance status (n=3), quality of life (QoL) (n=1) and return to work (n=6). The proportion of patients who returned to work within 12 months of surgery was 84% (95% confidence interval [CI] 50-96%, I-squared=38%, 5 studies) for gross total resection, 66% (95% CI 14-96%, I^2^=57%, 5 studies) for subtotal resection, and 31% (95% CI 4-82%, I^2^= 0%, 4 studies) for partial resection. There was insufficient data on other functional outcomes for quantitative synthesis.

**Conclusion:** A higher proportion of DLGG patients returned to work following gross total resection compared with those who had a subtotal or partial resection. Further studies with standardized assessments can clarify the association between EoR and different functional outcomes.

**Importance of the Study:** The association between lower residual tumor volume and better survival in people with diffuse low-grade glioma (DLGG) has driven surgical research into maximizing resection. However, innovative interventions should be evaluated against both oncological outcomes and functional outcomes. This systematic review included all studies reporting post-operative functional outcomes stratified by the extent of resection in people aged ≥18 years with DLGG. Two studies reported neurocognition, three reported performance status, one reported quality of life, and six reported return-to-work. Our meta-analysis demonstrated a higher proportion of patients returning to work within 12 months for those who had gross total resection (84%) compared to subtotal (66%) and partial (31%) resections. Our results suggest that increased EoR may be associated with return-to-work, but direct comparative studies should verify this finding and could examine other important functional outcomes.

**Key Points:**

1. Seven studies reported functional outcomes stratified by EoR in DLGG patients.
2. Return-to-work (RTW) was the most reported functional outcome.
3. 84% undergoing GTR of DLGG RTW within 12 months compared to STR (66%) or PR (31%).

## Introduction

Diffuse low-grade glioma (DLGG) is ultimately an incurable disease.^1^ It has a predilection for cortical and subcortical white matter tracts fundamental for both motor and speech function.^2,3^ The essence of patient management is an individualized approach employing advanced surgical and oncological methods.^4^ Early and maximal surgical resection allows a histological diagnosis to be established and may influence adjuvant therapy. ^5–7^ The extent of resection (EoR), as assessed on volumetric MRI FLAIR sequences is a positive prognosticator on overall survival (OS) and time to tumor progression (TTP). ^8,9^ However, the oncological benefit must be balanced against the risk of functional deficit that lowers the quality of life (QoL).^10^

Functional outcomes are complex. These outcomes often refer to the ability to carry out activities of daily living and QoL. In people with DLGG, there are additional relevant outcomes. Seizures are common in people with DLGG. Control of seizure, while being a clinical outcome, can have a profound impact on patients’ daily living. Both tumor and cranial surgery can cause cognitive impairments, rendering neurocognition and neuropsychology important functional outcomes. Advances in surgical techniques such as awake surgery and cognitive mapping aim to preserve cognitive functions while enabling maximal resection. Evaluation of modern neurosurgical techniques and novel therapies should include clinical and functional outcomes.

The surgical objective is maximal safe resection, which is associated with better oncological outcomes in DLGG.^11^ However, there is uncertainty about the relationship between EoR and functional outcome. This systematic review summarized functional outcomes after surgery for DLGG stratified by EoR.

## Materials and Methods

This systematic review was registered in PROSPERO (CRD42021238387) and reported according to Preferred Reporting Items for Systematic Reviews and Meta-Analyses (PRISMA) guidelines.

### Eligibility criteria

This systematic review included all studies of patients aged ≥18 years with histologically diagnosed supratentorial DLGG undergoing tumor resection with documented post-operative functional outcome stratified by EoR. Studies featuring any additional interventions not directly affecting EoR were excluded because of the likely bias in patient selection and confounding intervention effects. Exclusion criteria included patients aged <18 years, high grade glioma, previously treated DLGG and studies that did not provide comparative data on outcomes of interest or otherwise did not meet inclusion criteria. Studies with a sample size of <10 patients were excluded owing to a potential risk of publication bias.

### Information sources and search strategy

MEDLINE, EMBASE and the Cochrane Central Register of Controlled Trials (CENTRAL) were searched on 19/02/2021. The full search strategy is in supplementary materials (Tables 1.1, 1.2 & 1.3). Retrieved search results were combined and uploaded to Covidence – an online web-based□platform for systematic review record-keeping and management.^12^

### Selection process and data collection

Two reviewers (ME & EB) independently screened each article. Discrepancies during title/abstract screening and full-text eligibility assessment were resolved by discussion between both reviewers and with a senior reviewer (MTCP). Two independent reviewers (ME & EB) extracted data from each included study. Uncertainties were resolved with a senior reviewer (MTCP). For studies that reporting functional outcomes after DLGG surgery but not stratified by EoR, we contacted the corresponding author to seek clarification about the missing or uncertain data. If the corresponding author did not respond after three weeks, inclusion and data extraction decisions would be made in accordance with the available published material.

### Data Items

We collected data on study characteristics, extent of resection and post-operative functional outcomes. Study characteristics included year of publication, country of primary affiliation, study design, duration of patient recruitment and total number of patients recruited. For EoR, we collected the author defined EoR. We recorded both how and when EoR was ascertained. Resections were classified into the following groups: partial resection (PR), subtotal resection (STR), gross total resection (GTR) and supratotal resection (SpTR) according to author-defined subcategories on EoR.

Any objective measurement of a pre-defined postoperative functional outcome stratified by EoR was recorded. Pre-defined post-operative functional outcomes were as follows: performance status, quality of life, neurocognition and time taken to return to work. All included studies were comprehensively searched for time points of outcome acquisition and instruments used to measure each individual outcome (supplementary materials).

### Risk of Bias Assessment

We based our risk of bias assessment on the Newcastle-Ottawa Scale (NOS) for observational studies.^13^ The risk of bias within each individual study was independently assessed at the time of data extraction. A□senior□reviewer□was delegated for mediation in the event of disagreement that could not be resolved following discussion between reviewers. Certainty assessment was not applicable to this review. We used funnel plots and Egger’s test to assess publication bias.

### Synthesis Methods & Statistical Analysis

We identified studies of moderate to high quality (NOS >6) for meta-analysis. Where there were four or more studies reporting the same functional outcome at a uniform timepoint, results were summarized in our meta-analysis. If the proportion of patients was reported, then we meta-analyzed the proportions per EoR groups. We planned not to meta-analyze effect size because of anticipated study design heterogeneity. Results for studies included in qualitative analysis were presented in tables. Forest plots were used to illustrate the results of studies included in meta-analysis. For each outcome of interest, the proportion and its respective 95% confidence interval (CI) were calculated using a DerSimonian-Laird random effects model. We used I-squared (I^2^) and Tau-squared (τ^2^) statistics to assess heterogeneity. No subgroup analyses were planned to explore study heterogeneity. No sensitivity analyses were planned to assess the robustness of the synthesized results. We used R software version 4.1.2 using packages ‘*meta’* (version 5.5-0) and ‘metafor’ (version 3.4-0) for our analyses.^14^

## Results

There were 1,183 records retrieved from our search, of which seven eligible studies of 234 adults with DLGG reporting functional outcome after surgical resection were included in this review.^15–20^ Study demographics and characteristics are shown (Table 1). One study reported two independent cohorts in one study with outcome separately available for each cohort.^18^ The overall risk of bias as measured by the Newcastle-Ottawa Scale in all included studies was low (Table 2).

**Table 1.** Observational studies of post-operative functional outcomes stratified by extent of resection in patients with diffuse low-grade glioma

| Study |  | Ng et al., 2019 | Muto et al., 2018 | Lima & Duffau, 2015 | Moritz-Gasser et al., 2012 (Study 2) | Moritz-Gasser et al., 2012 (Study 1) | Duffau et al., 2009 | Budrukkar et al., 2009 |
| --- | --- | --- | --- | --- | --- | --- | --- | --- |
| Design |  | Prospective | Prospective | Prospective | Prospective | Retrospective | Retrospective | Prospective |
| Country |  | France | France | France | France | France | France | India |
| Recruitment period |  | December 1998 to November 2017 | March 2010 to March 2016 | December 1998 and September 2012 | 2009 to 2010 | Before 2008 | October 1997 to October 2008 | 1st January to 31 December 2008 |
| Sample size (n) |  | 74 | 39 | 14 | 12 | 11 | 13 | 71 |
| Mean Age (± SD) | | 35.7 (± 9.7) | 38 (± 12) | 38 (± 11) | 38 (± 11) | 38 (± 5) | 35 (± 12) | - |
| Gender (Male %) |  | 41.90% | 46% | 36% | 71% | 55% | 54% | 67% |
| Extent of Resection (EOR) | Partial | - | 12.80% | - | 16.70% | 45.50% | 15.30% | - |
|  | Sub-Total Resection (STR) | 41.90% | 41% | - | 58.30% | 54.50% | 69.20% | - |
|  | Gross Total Resection (GTR) | 29.70% | 35.90% | 71.40% | 25% | - | 15.30% | - |
|  | Supratotal Resection (SpTR) | 28.40% | 10.30% | 28.60% | - | - | - | - |
| Objective assessment of EOR? |  | Yes | No | Yes | Yes | Yes | No | No |
| Functional Outcomes |  | PS; RTW | RTW | PS; RTW | Ncog; RTW | Ncog; RTW | PS; RTW | QoL |

**Table 2.** Overall Newcastle-Ottawa score & quality assessment scores

| **Study** | **Quality**<br>**Assessment:**<br>**Selection**<br>**Score** | **Quality**<br>**Assessment:**<br>**Comparability**<br>**Score** | **Quality**<br>**Assessment:**<br>**Outcome**<br>**Score** | **Quality**<br>**Assessment:**<br>**Total Score** | **Quality**<br>**Classification:**<br>**(Poor/Fair/Good)** | **Risk of Bias**<br>**(Low/High/Very**<br>**High)** |
| --- | --- | --- | --- | --- | --- | --- |
| **Ng et al., 2019** | **** | ** | *** | 9 | Good | Low |
| **Muto et al., 2018** | **** | ** | *** | 9 | Good | Low |
| **Lima & Duffau, 2015** | **** | ** | *** | 9 | Good | Low |
| **Moritz-Gasser et al., 2012 (Study 1)** | *** | ** | *** | 8 | Good | Low |
| **Moritz-Gasser et al., 2012 (Study 2)** | *** | ** | *** | 8 | Good | Low |
| **Duffau et al., 2009** | **** | ** | *** | 9 | Good | Low |
| **Budrukkar et al., 2009** | *** | * | ** | 6 | Good | Low |
| **Mean** | **3.6** | **1.9** | **2.9** | **8.3** | **Good** | **Low** |

Of the 234 adults with DLGG included, 146 adults were symptomatic and 88 had an incidental diagnosis. The mean age ranged from 35.7 to 38 years. The ratio of males to females in all studies was 1:0.89. All except one study were conducted in France.^15–18,20^ Five studies had a prospective design. ^16–20^ All studies were single-center studies. In 4 of the 7 included studies, EoR was objectively measured on MRI between 48 hours and 3 months post operatively. ^15–17,20^ The remaining three studies did not specify how exactly EoR was measured. ^18–20^ EoR was reported in cm³ (volumetric analysis) or according to the Berger and Berger-Yordanova classifications. ^18,21,22^ The proportion of surgical resections undertaken across all studies was as follows: 14 partial resections (8.6%), 69 sub-total resections (42.3%), 51 gross total resections (31.3%) and 29 supratotal resections (17.8%). The remaining 71 patients came from a final study which did not specify exact resection proportions for the cohort but did include post-operative QoL data stratified by EoR.^19^

Only one study measured post-operative QoL data stratified by EoR in patients undergoing surgery for DLGG.^19^ Three studies recorded individual post-operative Karnofsky performance status (KPS) at three months alongside corresponding EoR data. ^15–17^ Two studies provided individual post-operative neurocognitive assessment data.^18^ 6 out of 7 studies reported post-operative return-to-work (RTW). ^15–18,20^

### Quality of Life

One study reported QoL following resection of DLGG by EoR.^19^ This was a single-center prospective study in India that included 71 consecutive adults with DLGG and reported post-operative QoL before starting adjuvant oncological treatment. Post-operative QoL was evaluated using the European Organization for Research and Treatment of Cancer (EORTC) Quality of Life of Cancer Patients (QLQ-C30) and Brain Cancer Module (QLQ-BN20) questionnaires.^23^ The mean EORTC QLQ-C30 global score for all post-operative DLGG patients was 61.9±33.8. For those undergoing partial resection and total excision of DLGG, the mean EORTC QLQ-C30 global score was 66.7 and 58.3 respectively. The type of surgery (biopsy vs. complete resection) had no influence on QoL score in univariable analysis (p=0.284).

### Performance Status

One retrospective case series in France included 24 patients who each underwent awake surgery with intraoperative direct electrical mapping for an insular WHO grade II glioma involving dominant hemisphere.^15^ KPS score was evaluated for each patient before surgery. 13 patients did not receive adjuvant treatment at 3 months post op. For the PR group (n=2), KPS at 3 months after surgery had improved in both patients. For those patients who had a STR (n=9), all but one had stable or improved KPS at 3 months. Both patients who underwent GTR (n=2) had improved KPS at 3 months post-operatively.

Another study in France reported a prospective series of 21 patients with incidental DLGG who underwent awake surgery with intraoperative direct electrical mapping for cortical and subcortical eloquent structures.^16^ The minimum follow-up duration was 20 months. KPS was evaluated 3 months after surgery. Three months after surgery, all 21 patients’ KPS remained unchanged at 100 regardless of extent of resection.

A third study in France reported a single-center series of 74 patients with incidental DLGG who underwent awake surgery with intraoperative functional mapping through cortical and subcortical direct electrostimulation.^20^ Post-operative clinical and radiological data including EoR were prospectively collected. KPS was recorded pre-operatively and at 3 months post-operatively. KPS remained unchanged at 100 in all patients regardless of EoR. The minimum follow-up period for all patients was 12 months.

### Neurocognition

Only one study reported post-operative neurocognition in patients with DLGG who underwent awake surgery with intraoperative brain mapping.^18^ The study ‘s primary objective was to determine if lexical access was correlated with post-operative return to work in DLGG patients. This study consisted of two separate case-control series.

The first retrospective case-control series featured 11 patients who underwent post-operative assessment via a battery of neurocognitive tests. 5 patients had a partial resection whilst 6 patients had a subtotal resection. For language assessment, the picture naming ‘‘DO.80 test’’ was used.^24^ This test has two components: naming time (NT) and naming accuracy (NA). The mean NA score was 0.38 for those undergoing PR and 0.69 for the STR group. The mean NT score was -9.22 for those undergoing PR and -6.97 in the STR group.

A second prospective case-control series featured 12 patients who performed pre- and post-operative language assessment, using the picture naming ‘‘DO.80 test’’ and had a median follow-up of 9 months. 2 patients had undergone a partial resection, 7 patients had a subtotal resection, and 3 patients underwent gross total resection. Mean NA scores were 0.21, -2.38 and -1.09 for the PR, STR and GTR groups respectively. Mean NT scores were -1.74, -2.69 and -2.91 for the PR, STR and GTR groups respectively.

### Return to Work

Of the seven included studies, six studies reported post-operative RTW data. ^15–18,20^ Meta-analyses of the proportion of patients who returned to work within 12 months of an operation for DLGG was conducted for each subgroup of EoR namely, PR, STR, GTR & SpTR. The proportion of patients who returned to work within 12 months of surgery was 84% (95% confidence interval [CI] 50-96%, I-squared=38%, 5 studies) for gross total resection, 66% (95% CI 14-96%, I-squared=57%, 5 studies) for subtotal resection, and 31% (95% CI 4-82%, I-squared 0%, 4 studies) for partial resection. (Figure 2). The proportion of patients who returned to work within 12 months was 69% (95% CI 50-83%, I-squared 0%, 3 studies) in the supratotal resection group. All funnel plots constructed to assess for publication bias across each meta-analysis were symmetric. (Supplementary Material Figures 3.1, 3.2, 3.3 & 3.4)

**Figure 1.**
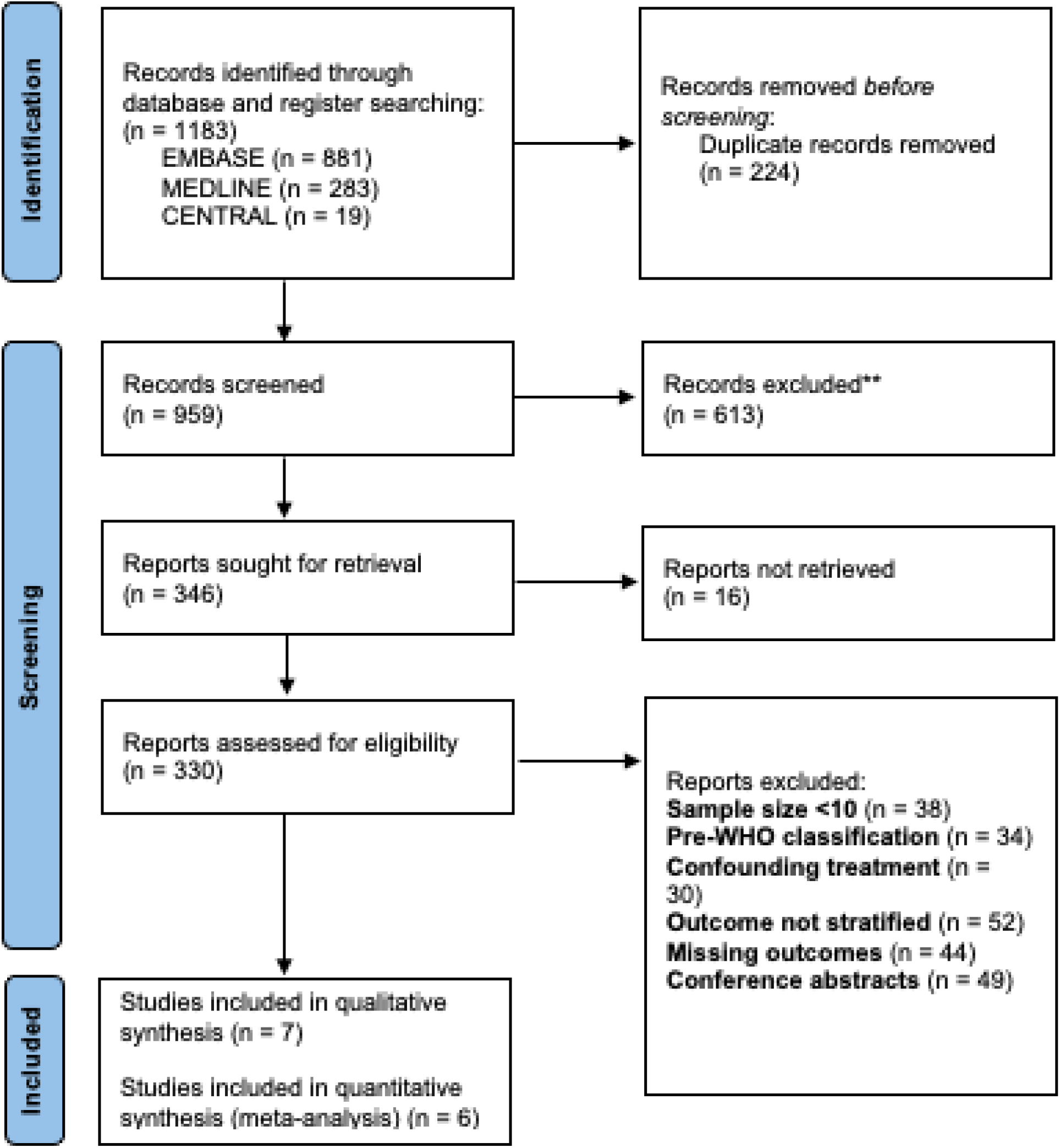
Flow chart of database searching strategy detailing the report selection procedure as outlined in the Preferred Reporting Items for Systematic Reviews and Meta-Analyses 2020 guideline

**Figure 2.**
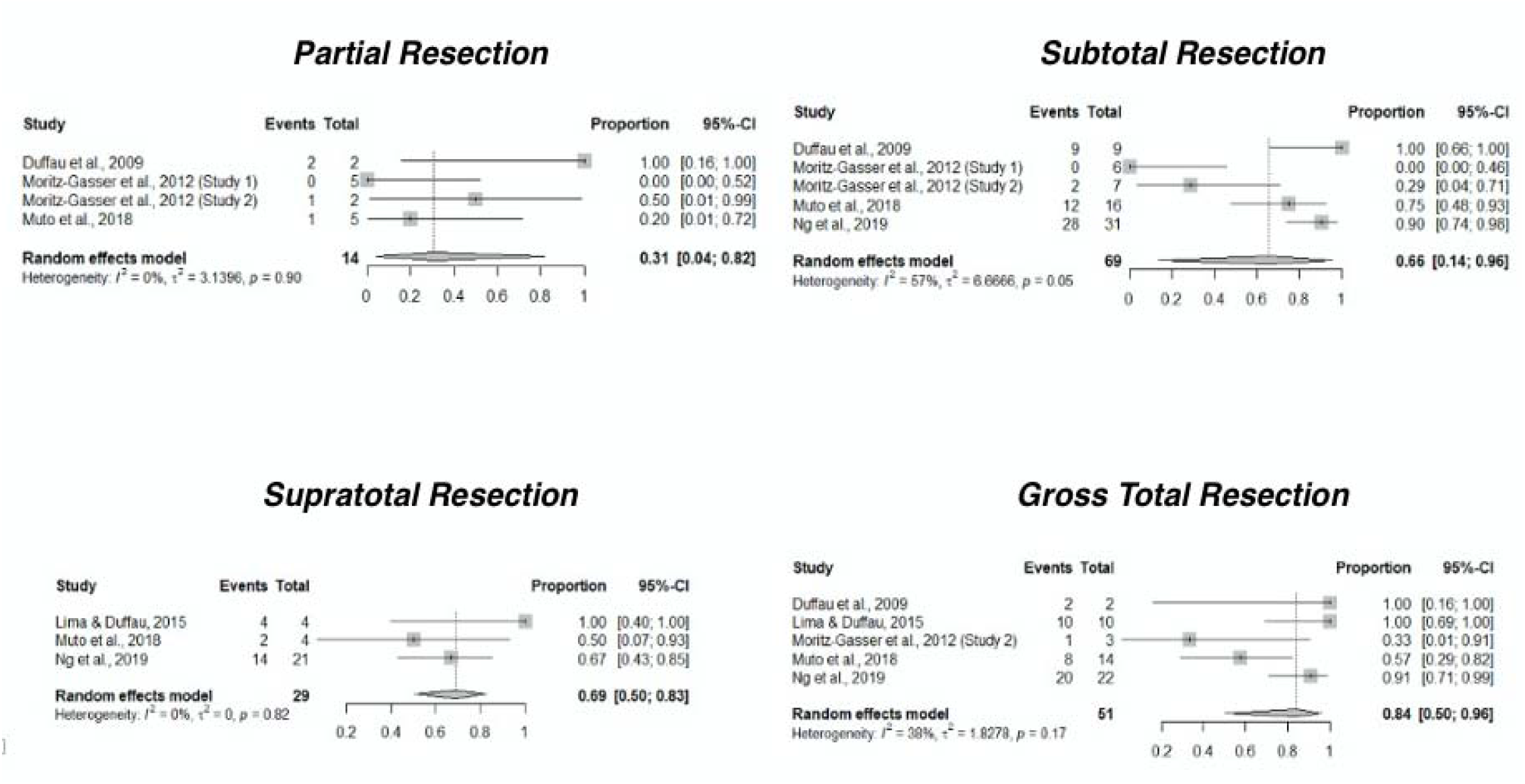
Meta-analyses of proportions of patients who returned to work within 12 months following partial resection, subtotal resection, gross total resection and supratotal resection of diffuse low-grade glioma

## Discussion

Only seven studies reported functional outcomes stratified by EoR in people who had surgical resection for DLGG. Individual studies reported sustained or improved functional outcomes following complete resection, though direct comparison cohorts were lacking. Meta-analysis of six cohorts suggested higher proportion of patients returned to work in those with GTR.

### Return to work

The pooled proportion estimate of people returning to work within one year was higher for people who had GTR at 84% compared to those who had STR and PR at 66% and 31%, respectively (Figure 2). This finding is supportive of the current strategy of maximal safe resection. However, tumor location and tumor volume are likely confounders for the association between EoR and functional outcome. It is difficult to confirm the association between EoR and return-to-work without direct comparative and adjusted analysis. EoR is also a proxy outcome for people returning to a level of functional status that allows them to return to work. This outcome depends on the physical, cognitive, and mental demands of the occupation. The heterogeneous occupations can also introduce a degree of outcome measurement error. These limitations make the interpretation and clinical translation of results difficult. Objective and standardized measures of functional and neurocognitive status may better inform the association between EoR and functional outcome.

### Other functional outcomes

Some studies in this review reported quality of life, performance status and neurocognition in relation to the EoR. Data was insufficient for evidence synthesis by meta-analysis. This lack of data is consistent with another systematic review of functional outcomes after glioma surgery.^25^ The systematic review included 160 studies and only 8% and 6% studies reported neurocognition and health reported quality of life, respectively. The authors have initiated a survey of current practices to create a consensus on a standard set of assessments and reporting guidelines.^25^ The results are pending especially as the optimal timing of measuring outcomes and battery of tests for each functional outcome domain is unknown.

Outcome ascertainment for functional outcomes is also challenging. While a consensus can agree on the assessment tools, observer bias can affect consistency of assessment results. Patient-reported outcomes are generally regarded as more clinically relevant. However, in patients who may not be able to complete an assessment themselves, family or friends may help answer questions though can introduce a degree of measurement error. As such, there are categories of outcomes: patient-reported outcomes, clinician-reported outcomes, observer-reported outcomes, and task-based outcomes. The optimal functional outcome assessment should incorporate elements from all these categories informed by public and patient involvement.

### Strengths and limitations

This systematic review provided a benchmark of functional outcomes stratified by EoR following resective surgery for DLGG. Findings can inform study design of future clinical studies. We were able to meta-analyze results from six moderate to high quality studies to provide estimates of return-to-work within one year.

Only return-to-work had sufficient studies to enter meta-analysis. We were not able to summarize other functional outcomes. Taking the absolute numbers for meta-analyses rather than the adjusted effect size from studies make our analyses prone to the effects of confounding. However, none of these studies provided adjusted effect sizes. Our results here provide the best estimate available based on existing evidence. With the updates of the 2021 WHO classification of brain tumors, findings in this review may not apply to all the molecularly defined low-grade gliomas. Futures studies need to report detailed clinical features and molecular characteristics to be relevant.

### Conclusion

Functional ability is an important outcome alongside progression-free and overall survival. This systematic review showed that few studies have directly assessed the association between EoR and functional outcomes. Though there was a suggestion that higher EoR is associated with higher chance of returning to work within 12 months, but direct comparative studies were lacking. Future studies evaluating novel surgical strategies should incorporate a clinically relevant comparison group and prospectively assess traditional outcomes and functional abilities.

## Supporting information

PRISMA 2020 Checklist

## Data Availability

All data produced in the present study are available upon reasonable request to the authors

## Acknowledgements

Findings from this review were presented at the XVII World Congress of Neurosurgery (WFNS 2022) Meeting held in Bogotá, Colombia from the 13th to the 18th of March 2022.

