## Supplementary Material for "Association of Extent of Resection and Functional Outcomes in Diffuse Low-Grade Glioma: Systematic Review & Meta-Analysis"

| 1 | ((diffuse astrocytoma) OR oligodendroglioma OR (pleomorphic xanthoastrocytoma) OR (chordoid glioma of third ventricle)).mp |
| --- | --- |
| 2 | ((grade adj2 II) OR (grade adj2 2) OR (low adj2 grade) OR low-grade OR diffuse).mp AND (glioma* OR astrocytoma* OR oligoastrocytoma*).mp |
| 3 | surgery.fs OR exp surgical procedures, operative/ OR exp neurosurgery/ OR ((surg* adj2 resec*) OR craniotomy* OR debulk* OR excis* OR resec*).mp |
| 4 | ((extent adj3 of adj3 resection) OR eor OR supratotal OR (gross adj3 total) OR gross-total OR sub?total).mp |
| 5 | exp quality of life/ OR ((quality adj4 life) OR QoL OR HRQoL OR (health adj3 related adj3 quality) OR (health-related adj3 quality) OR (functional adj3 outcome) OR (EORTC adj2 QLQ-BN20) OR (EORTC adj2 QLQ-C30) OR SF?36 OR FACT-Br OR WHO?QOL* OR EuroQoL OR EuroQol-5D OR EQ?5D* OR SEIQoL* OR SQoL OR SQLP OR (Sintonen adj2 15D) OR NHP).mp OR exp return to work/ OR exp neuropsychological test/ OR exp cognitive dysfunction/ |
| 6 | (1 OR 2) AND (3 OR 4) AND 5 |
| 7 | Review/ OR Comment/ OR Editorial/ |
| 8 | 6 NOT 7 |

*Table 1.1 - MEDLINE Search Strategy*

| 1 | ((diffuse astrocytoma) OR oligodendroglioma OR (pleomorphic xanthoastrocytoma) OR (chordoid glioma of third ventricle)).mp |
| --- | --- |
| 2 | ((grade adj2 II) OR (grade adj2 2) OR (low adj2 grade) OR low-grade OR diffuse).mp AND (glioma* OR astrocytoma* OR oligoastrocytoma*).mp |
| 3 | surgery.fs OR exp surgical procedures, operative/ OR exp neurosurgery/ OR ((surg* adj2 resec*) OR craniotomy* OR debulk* OR excis* OR resec*).mp |
| 4 | ((extent adj3 of adj3 resection) OR eor OR supratotal OR (gross adj3 total) OR gross-total OR sub?total).mp |
| 5 | exp quality of life/ OR ((quality adj4 life) OR QoL OR HRQoL OR (health adj3 related adj3 quality) OR (health-related adj3 quality) OR (functional adj3 outcome) OR (EORTC adj2 QLQ-BN20) OR (EORTC adj2 QLQ-C30) OR SF?36 OR FACT-Br OR WHO?QOL* OR EuroQoL OR EuroQol-5D OR EQ?5D* OR SEIQoL* OR SQoL OR SQLP OR (Sintonen adj2 15D) OR NHP).mp OR exp return to work/ OR exp Work resumption/ OR exp neuropsychological test/ OR exp cognitive defect/ |
| 6 | (1 OR 2) AND (3 OR 4) AND 5 |
| 7 | Review/ OR Comment/ OR Editorial/ |
| 8 | 6 NOT 7 |

*Table 1.2 - EMBASE Search Strategy*

| 1 | ((diffuse astrocytoma) OR oligodendroglioma OR (pleomorphic xanthoastrocytoma) OR (chordoid glioma of third ventricle)) |
| --- | --- |
| 2 | ((grade adj2 II) OR (grade adj2 2) OR (low adj2 grade) OR low-grade OR diffuse) AND (glioma* OR astrocytoma* OR oligoastrocytoma*) |
| 3 | surgery.fs OR ((surg* adj2 resec*) OR craniotomy* OR debulk* OR excis* OR resec* |
| 4 | MeSH descriptor: [Surgical Procedures, Operative] explode all trees |
| 5 | MeSH descriptor: [Neurosurgery] explode all trees |
| 6 | ((extent adj3 of adj3 resection) OR eor OR supratotal OR (gross adj3 total) OR gross-total OR sub?total).mp |
| 7 | MeSH descriptor: [Quality of Life] explode all trees |
| 8 | ((quality adj4 life) OR QoL OR HRQoL OR (health adj3 related adj3 quality) OR (health-related adj3 quality) OR (functional adj3 outcome) OR (EORTC adj2 QLQ-BN20) OR (EORTC adj2 QLQ-C30) OR SF?36 OR FACT-Br OR WHO?QOL* OR EuroQoL OR EuroQol-5D OR EQ?5D* OR SEIQoL* OR SQoL OR SQLP OR (Sintonen adj2 15D) OR NHP).mp |
| 9 | MeSH descriptor: [Return to Work] explode all trees |
| 10 | MeSH descriptor: [Neuropsychological Tests] explode all trees |
| 11 | MeSH descriptor: [Cognitive Dysfunction] explode all trees |
| 12 | (#1 OR #2) AND (#3 OR #4 OR #5 OR #6) AND (#7 OR #8 OR #9 OR #10 OR #11) |

*Table 1.3 - CENTRAL Search Strategy*

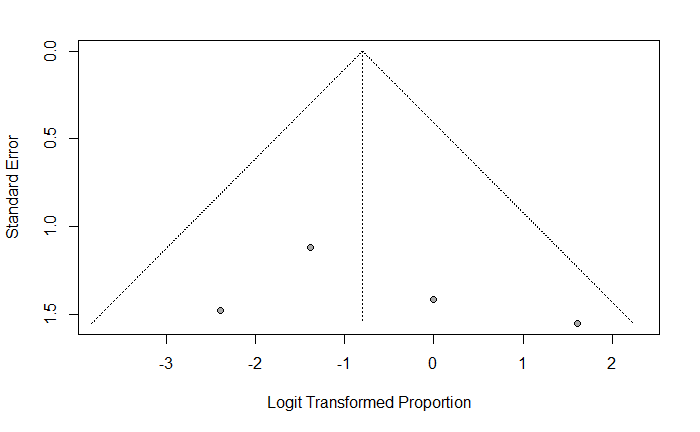
*Figure 3.1 - Funnel plot depiction of meta-analysis of proportion of patients who returned to work within 12 months following partial resection of DLGG*

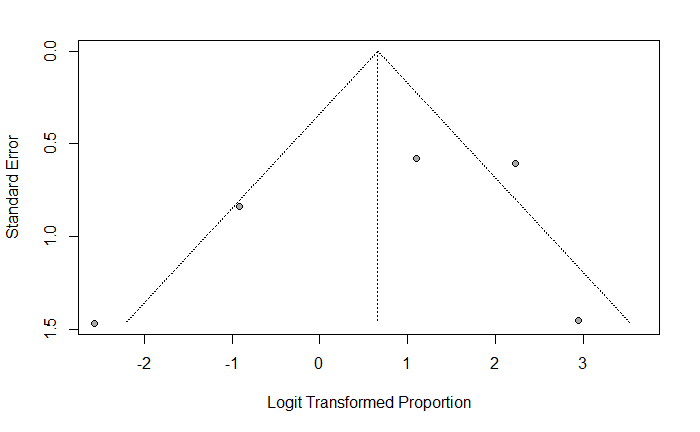
*Figure 3.2 - Funnel plot depiction of meta-analysis of proportion of patients who returned to work within 12 months following sub-total resection of DLGG*

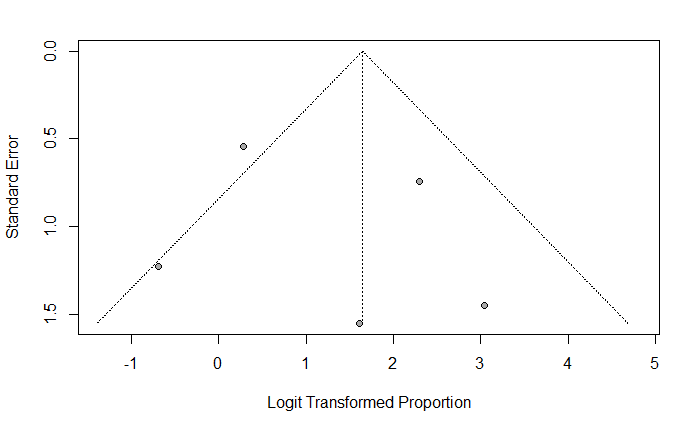
*Figure 3.3 - Funnel plot depiction of meta-analysis of proportion of patients who returned to work within 12 months following gross total resection of DLGG*

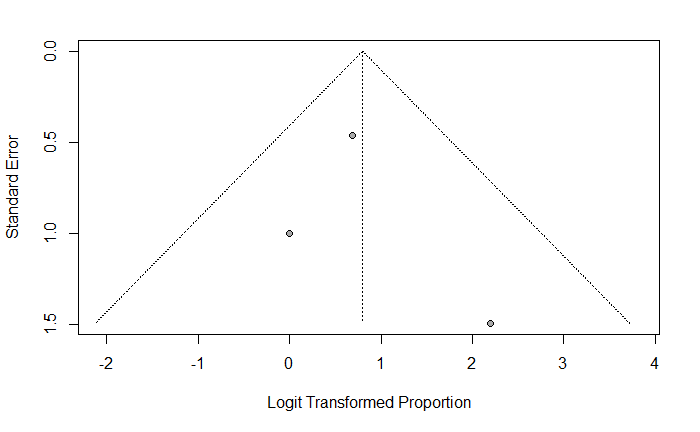

*Figure 3.4 – Funnel plot depiction of meta-analysis of proportion of patients who returned to work within 12 months following supratotal resection of DLGG*
